# Gaps in HIV testing for children of mothers with known HIV positive status: Results from Population-based HIV Impact Assessments (PHIA) in Sub-Saharan Africa (2015-2019)

**DOI:** 10.64898/2026.03.19.26348854

**Authors:** Lennah Nyabiage, Susan Gachau, Sasi Jonnalagadda, Sileshi Lulseged, Dumbani Kayira, Alinune Nathanael Kabaghe, Ismelda Kutara, Sabin Nsanzimana, Veronicah Mugisha, Marie Louise Umwagange, Khumbo Namachapa, Edward Machage, Jonathan M. Grund, George Mgomella, Megumi Itoh, Talent Maphosa, Owen Mugurungi, Wondimu Teferi, J. Danielle Sharpe, Hannah M. Canepa, Mary Mahy, Jessica Gross, Andrew C. Voetsch

## Abstract

**Introduction:** HIV testing for children of women living with HIV (WLHIV) is an efficient method of diagnosing HIV in children. We analyzed pooled data from 13 Population-based HIV Impact Assessments (PHIA) conducted from 2015 through 2019 to determine the gap in diagnosing HIV in children of WLHIV.

**Methods:** In each PHIA, children younger than 15 years in a subset of households were sampled for HIV testing. Mother-reported responses on child’s status were linked to maternal interviews and biomarker data. Analysis was restricted to children whose mothers were alive, older than 15 years and aware of their HIV-positive status prior to the survey. We calculated weighted proportions of children who were never previously tested and proportion of children living with HIV (CLHIV) with no evidence of antiretroviral treatment (ART) use (categorized as newly diagnosed). Survey weights were pooled across all PHIAs to account for survey design and nonresponse.

**Results:** Of 4,234 WLHIV, 3,436 were aware of their HIV status and had at least one child (n=6,173) for whom responses were obtained. Of the 6,173 children, 43.5% (n=2,371) were reported as never been tested. Overall, 5,500 children provided blood for HIV testing during the survey. Newly diagnosed test positivity was 1.7% (90/5,191); 2.9% (61/2,120) among those with reported unknown HIV status and 0.9% (29/3,071) among those with reported HIV negative status. Among children with reported HIV positive status, 94.5% were confirmed by survey testing and of these, 91% had antiretrovirals (ARVs) detected.

**Conclusions:** Over 40% of children of WLHIV who were aware of their HIV positive status had never been tested for HIV. HIV positivity ranged between 0.9% to 2.9% while 9.0% of children known to be HIV positive were not on ART. The study calls for renewed efforts to enhance testing of children and treatment linkage for those diagnosed with HIV.

## Introduction

The Joint United Nations Programme on HIV/AIDS (UNAIDS) estimated that in 2024, 1.4 million children worldwide under the age of 15 were living with HIV, with 86% of them located in sub-Saharan Africa. In the same year, 120,000 children were newly infected with HIV, and 83% of these new infections occurred in sub-Saharan Africa [1].

Overall, 46% of all children living with HIV (CLHIV) had not accessed life-saving antiretroviral treatment (ART) in sub-Saharan Africa, potentially due to undiagnosed HIV infections [1]. The testing of HIV-exposed children in prevention of vertical transmission (PVT) of HIV programs remains suboptimal. As a result, children with perinatally acquired HIV are often diagnosed later in childhood, which increases their risk of morbidity and mortality [2].

Children of adults living with HIV have a higher likelihood of testing HIV positive, as up to 65% of undiagnosed children have a family member already enrolled in an ART program [3]. Testing children of HIV-positive parents has led to a remarkable improvement in the identification of children missed by PVT programs [4]. Family-based pediatric index testing requires elicitation of all children of HIV-positive parents and timely, safe, and ethical index testing [5]. In countries with high HIV prevalence, index testing strategies have found previously undiagnosed CLHIV among parents receiving ART [6, 7]. However, index testing coverage remains suboptimal, ranging from 14 – 71% [8, 9]. As countries move toward HIV epidemic control, family-based pediatric index testing to identify the remaining undiagnosed children continues to be a priority [10].

We analyzed data from 13 Population-based HIV Impact Assessment (PHIA) surveys in sub-Saharan African countries conducted between 2015 and 2019 to determine the proportion of undiagnosed HIV among children of women living with HIV (WLHIV) who were aware of their own HIV positive status. We also evaluated factors associated with lack of HIV testing among these children.

## Methods

### Study procedures

We analyzed data from PHIA surveys conducted from 2015 through 2019 in Cameroon, Côte d’Ivoire, Eswatini, Ethiopia, Kenya, Lesotho, Malawi, Namibia, Rwanda, Tanzania, Uganda, Zambia, and Zimbabwe. PHIA surveys are cross-sectional household surveys designed to measure the impact of the national HIV response through HIV incidence, prevalence, and viral suppression. A representative sample of persons aged 15 years and older and children less than 15 years was selected using a two-stage cluster design [11]. Stage one involved sampling census-derived primary sampling units or enumeration areas from households, which were sampled in the second stage. In each eligible household, all household members aged 15 years and older were eligible to participate in the survey, while children less than 15 years were selected in a sub-sample of households. In Eswatini, Lesotho, Malawi, Namibia, Zambia and Zimbabwe, all children in half of the selected households were randomly selected to participate in the survey. In Cameroon, Kenya, Tanzania, and Uganda a third of households with eligible children were randomly selected for survey participation. Sampling in Côte d’Ivoire included children of women who were HIV positive, deceased mothers, mothers with unknown HIV status and a sample of HIV negative mothers. In Rwanda, children aged 10-14 years of age in all sampled households were included in the survey [12]

All survey procedures at both the household and individual levels were conducted only after obtaining informed consent. In each sampled household, the head of the household provided informed consent and responded to all household-level questions. Thereafter, individual informed consent was obtained from all eligible adults within the household and assent was equally obtained from children aged 10 – 14 yrs. A standard questionnaire was used to interview eligible household members to collect information on awareness of their HIV status and ART status among household members who self-reported as HIV-positive. For children, information on HIV testing history, HIV status, and ART status was obtained from a parent or guardian.

Biomarker testing was offered to children with parent/guardian consent. Participants older than 18 months were tested for HIV in the sampled household using each country’s national algorithm. HIV test results were returned on the same day in accordance with HIV testing guidelines for adults and children in each country. Participants who tested HIV-positive in the household underwent confirmatory testing at the central survey laboratory using the Geenius™ HIV ½ supplemental assay (Bio-Rad, Hercules, California, United States). Children ≤18 months of age were screened for HIV infection using Determine™ HIV-1/2 (Abbott Molecular Inc., Des Plaines, Illinois, United States) at the household and confirmed through virologic testing using HIV TNA PCR using Abbott Real-Time HIV-1 Qualitative Assay (Abbott Molecular, Wiesbaden, Germany). All participants with a confirmed HIV-positive test underwent viral load testing to quantify HIV RNA using the Abbott RealTime HIV-1 assay with plasma or using the optimized one-spot protocol for DBS HIV-1 VL (HIV RNA copies per mL) using the fully automated Abbott m2000 System (Abbott Molecular, Des Plaines, Illinois, United States). To determine recent exposure to ART, dry blood spots (DBS) were tested for the presence of antiretroviral (ARV) medications using high-resolution liquid chromatography coupled with tandem mass spectrometry at the University of Cape Town, South Africa. Specific ARVs tested for adults and children varied by country and were consistent with prevailing national treatment guidelines [13].

### Statistical analysis

The adult interview and biomarker datasets were linked to the child datasets and pooled across all 13 countries. Sample weights based on sampling probabilities by age and sex and adjusted for non-response and post-stratification using national population estimates from the survey year were pooled across countries [14].

We restricted the analysis to children of women aged ≥ 15 years who tested HIV-positive in the survey and self-reported their own HIV-positive status or did not self-report their own HIV-positive status but had ARVs detected. Children who tested HIV positive and had detectable ARVs were categorized as known HIV positive while those who did not have detectable ARVs were categorized as newly diagnosed. Among this subgroup of children, we described the outcome of prior HIV testing as reported by their parent/guardians: unknown HIV status or never tested for HIV, HIV-negative and HIV-positive. We determined parent and child-level factors associated with children with unknown HIV status, using a logistic regression model with log link (using the survey-weighted generalized linear model (svyglm) in the R version 4.2 *survey* package).

To extrapolate the sample estimates to the broader target population, we employed the *svytotal* function from the R survey package. This approach allowed us to generate population-level totals by incorporating survey weights and adjusting for the sampling design. Pooled Jackknife replicate weights were used to calculate the corresponding 95% confidence intervals for the estimated adjusted prevalence ratios. All analyses were conducted in R version 4.2 and all results presented are weighted.

### Ethical considerations

All PHIA surveys were approved by Institutional Review Boards in each country, Westat and Columbia University. This activity was reviewed by CDC, deemed not research, and was conducted consistent with applicable federal law and CDC policy. The surveys were funded by the U.S. President’s Plan for AIDS Relief (PEPFAR) through the U.S. Centers for Disease Control and prevention. (Supplementary Table 1).

## Results

Of 4,234 WLHIV, 3,856 women had 7,641 linked biological children. Among these women, 3,436 were aware of their HIV-positive status prior to the survey and 80.8% (6,173) of the children were linked to them. On average, the total number of children elicited per woman was approximately 2. Of the 6,173 children, 4.5% were reported to be living with HIV, 52.0% were reported to have been tested at least once and had a negative test result, and 43.5% were reported to have never been tested. Overall, 5,500 of the 6,173 children had blood samples collected and tested for HIV during the survey and 7.0% tested positive. Of 2,371 children with reported unknown HIV status, 2,139 had blood samples collected for HIV testing. Among these, 80 (4.9%) tested HIV positive of whom 39.0% had detectable ARVs and were categorized as known HIV positive and 61.0% without detectable ARVs were categorized as newly diagnosed with HIV. The proportion newly diagnosed with HIV was 2.9% (61/2,120). Of the 3,503 children with reported HIV negative status, 3,080 had blood samples collected for HIV testing. Among these, 38 (1.5%) tested HIV positive, of whom 32.6% had detectable ARVs and were categorized as known HIV positive and 67.4% without detectable ARVs were categorized as newly diagnosed with HIV. The proportion newly diagnosed with HIV was 0.9% (29/3,071). Of the 299 children with reported positive HIV status, 281 had blood samples collected for HIV testing. Of these, 12 (5.5%) children tested HIV negative. Of the 269 children who tested HIV positive, 9.0% did not have detectable ARVs. These were as categorized as previously diagnosed but were not on ART at the time of the survey. The maternal reported HIV status of the child, survey HIV test results, and ARV detection results are shown in Figure 1.

**Figure 1:**
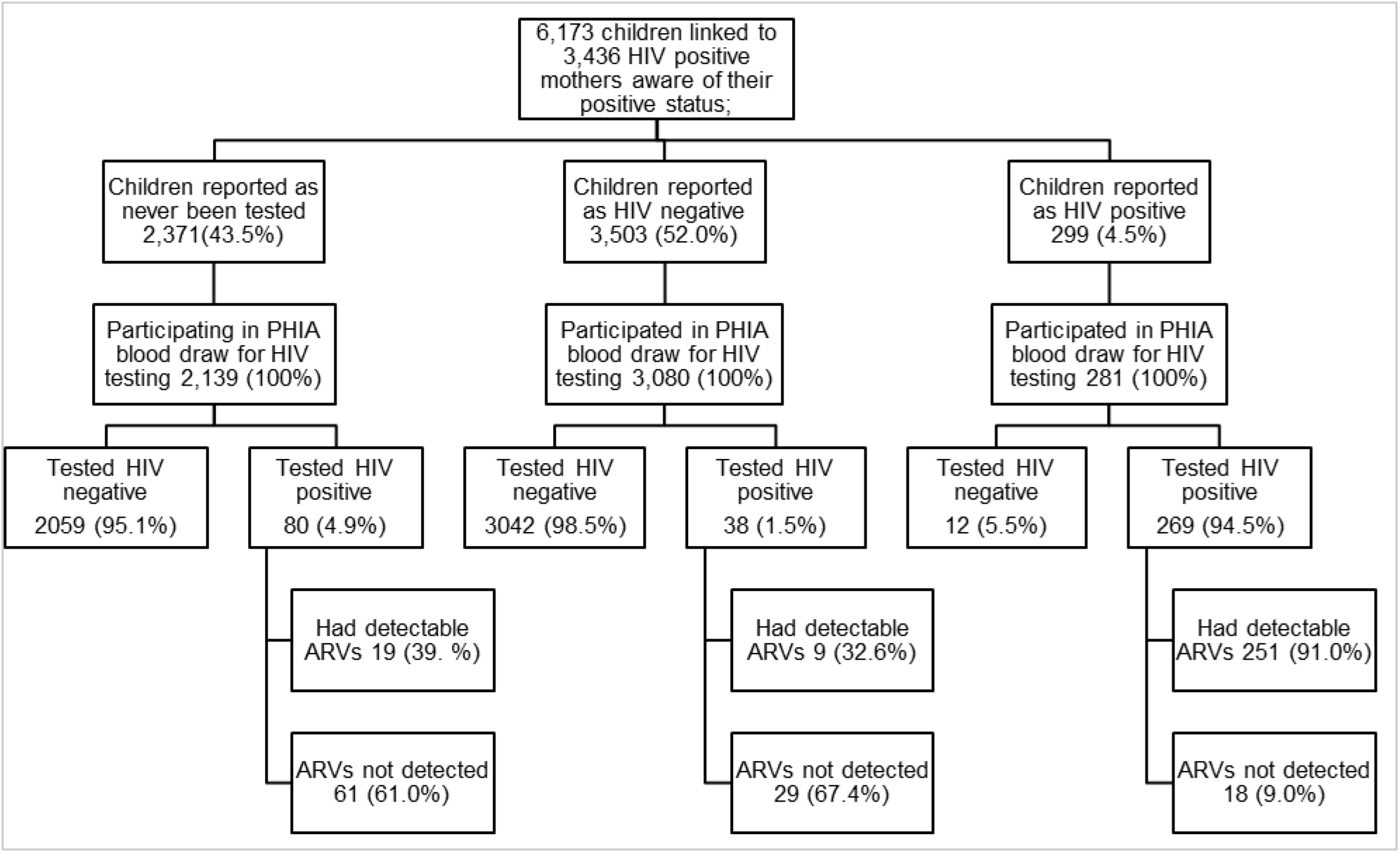
HIV testing outcomes for children of women living with HIV who were aware of their HIV positive status, Population-based HIV Impact Assessments, 2015-2019. \**Weights: Blood Test Replicate Weights*

Of the 2,371 children with reported unknown HIV status, 40.1% were between 10 and 14 years old, 64.7% were in school at the time of the survey, 89.0% had a mother who was on ART, 91.9% had a mother who was not sick in the last 12 months, and 94.3% had a father who was not sick in the past 12 months Of the 3,503 children reported to have tested HIV negative, 49.6% were under 5 years of age, 42.1% were too young to attend school, 94.0% had a mother taking ART, 92.9% had a mother who did not self-report as being sick in the last 12 months, and 93.0% had a father who had not been sick in the last 12 months (Table 1).

**Table 1:** Characteristics of children of women living with HIV who were aware of their own HIV positive status, by their parent/guardian reported HIV status, PHIA surveys conducted in 13 countries, 2015 – 2019.

| Characteristics | Children with unknown HIV status prior to the survey, n (weighted %), N = 2,371 | Children reported as HIV negative in last HIV test result prior to the survey, n (weighted %), N = 3,503 | Children reported as HIV positive in last HIV test result prior to the survey, n (weighted %), N = 299 |
| --- | --- | --- | --- |
| **Child characteristics** |  |  |  |
| Age (years) |  |  |  |
| <5 | 620 (27.1) | 1,595 (49.6) | 55 (25.7) |
| 5-9 | 770 (32.8) | 1,057 (27.7) | 112 (40.3) |
| 10-14 | 981 (40.1) | 851 (22.7) | 132 (34.0) |
| Child in school |  |  |  |
| Yes | 1,540 (64.7) | 1,899 (49.9) | 213 (58.4) |
| School age but not in school | 181 (7.8) | 298 (8.0) | 24 (11.1) |
| Not school age | 564 (27.5) | 1,303 (42.1) | 62 (30.5) |
| **Maternal characteristics** |  |  |  |
| Age (years) |  |  |  |
| <25 | 151 (6.0) | 255 (7.3) | 14 (7.5) |
| 25-29 | 375(16.2) | 635(17.4) | 35 (13.5) |
| 30-34 | 616(25.5) | 957 (26.5) | 72 (20.3) |
| ≥35 | 1,229 (52.4) | 1,656 (48.5) | 178 (58.7) |
| Marital status |  |  |  |
| Married | 931(52.9) | 1,483 (57.2) | 112 (54.1) |
| Living together | 178(12.3) | 265 (12.5) | 20 (7.0) |
| Widowed | 277 (15.1) | 416 (15.0) | 41(24.3) |
| Divorced | 103(10.7) | 115 (7.8) | 11 (11.9) |
| Separated | 100(9.0) | 197(7.5) | 12 (2.7) |
| Maternal education level |  |  |  |
| No primary | 206 (15.7) | 243(11.1) | 24 (13.2) |
| Primary | 991 (55.5) | 1,460 (54.1) | 131 (55.3) |
| Secondary | 588 (26.0) | 1,007 (31.5) | 67 (28.4) |
| College and more | 40 (2.8) | 91 (3.3) | 6 (3.1) |
| Maternal ART status |  |  |  |
| Mother on ART | 2,098 (89.0) | 3,276(94.0) | 281(91.4) |
| Mother not on ART | 257 (11.0) | 225 (6.0) | 18 (8.6) |
| Mother sick in the past 12 months |  |  |  |
| Yes | 197(8.1) | 289(7.1) | 32(8.9) |
| No | 2,167 (91.9) | 3,205 (92.9) | 267 (91.1) |
| Father sick in the past 12 months |  |  |  |
| Yes | 118(5.7) | 183(7.0) | 15(10.2) |
| No | 1,756 (94.3) | 2,612 (93.0) | 210 (89.8) |

\*Weights: Blood Test Replicate Weights

Factors significantly associated with a child never having been tested for HIV prior to the survey included living in a rural area, illness in one of the biological parents in the 12 months prior to the survey, not visiting a health care worker in the last 12 months, and having a child not enrolled in school. Maternal characteristics associated with a child never having been tested for HIV include not disclosing HIV status, not taking ART, not attending antenatal clinic (ANC) and not being HIV virally suppressed. (Table 2).

**Table 2:**
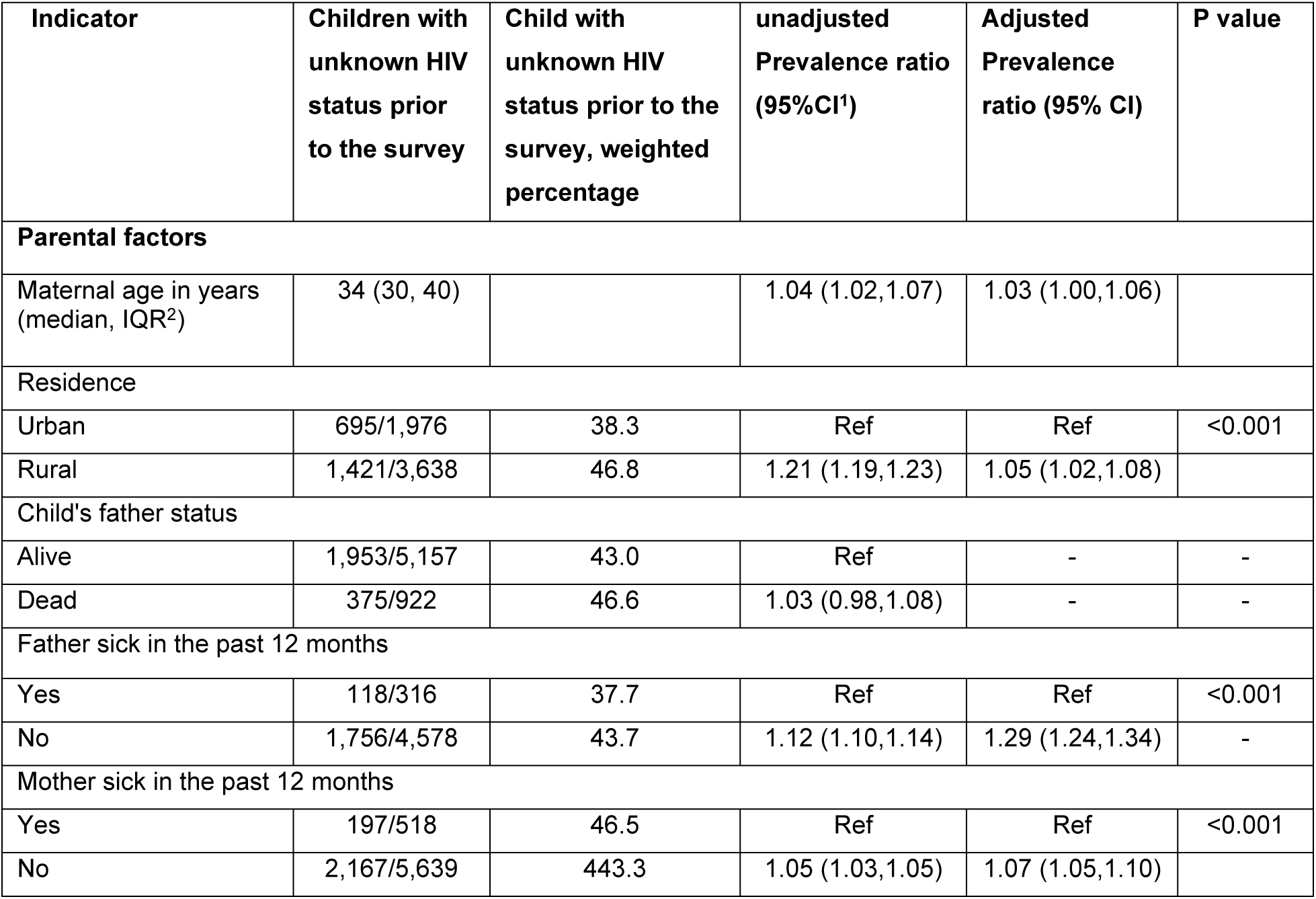
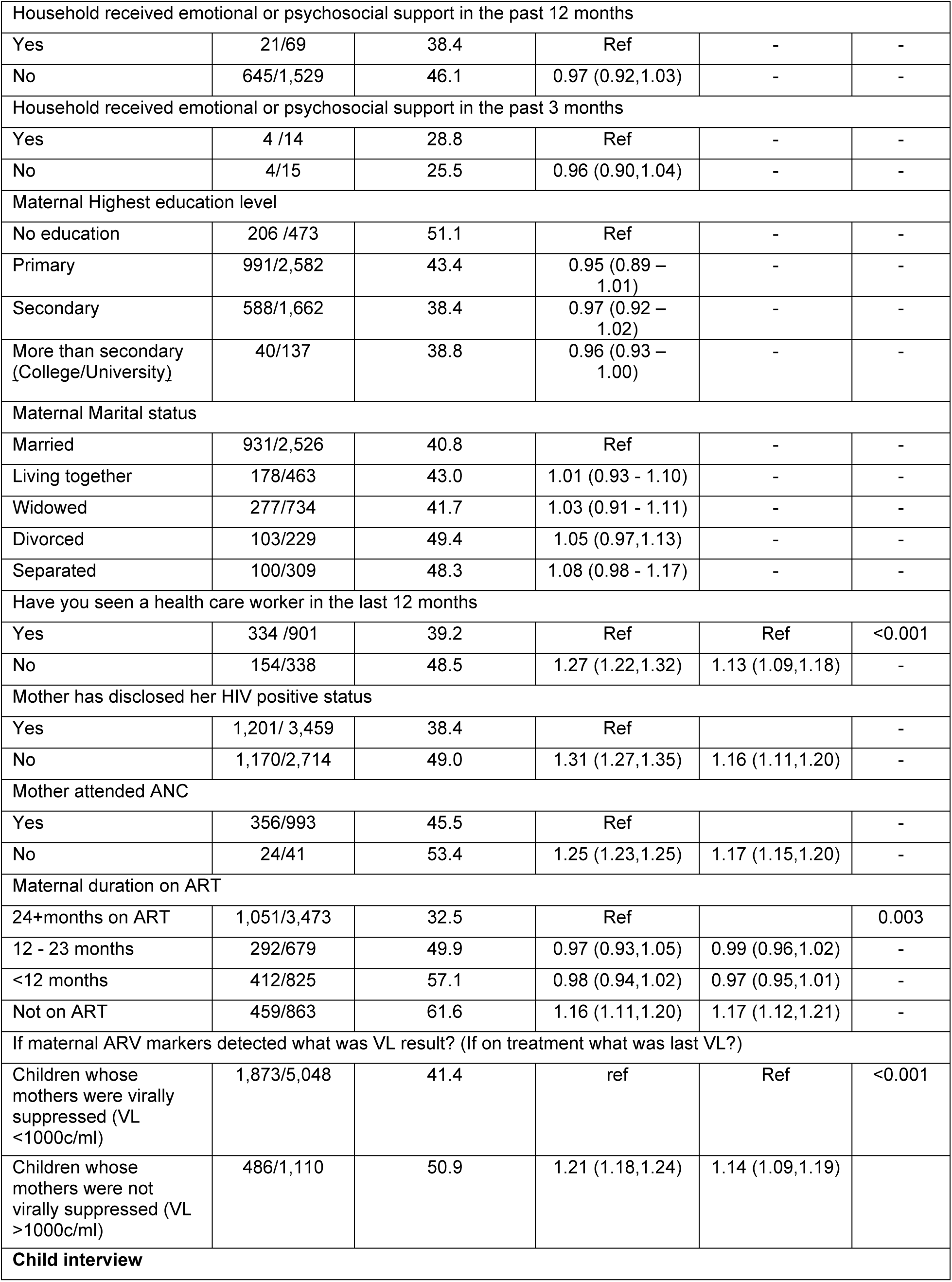
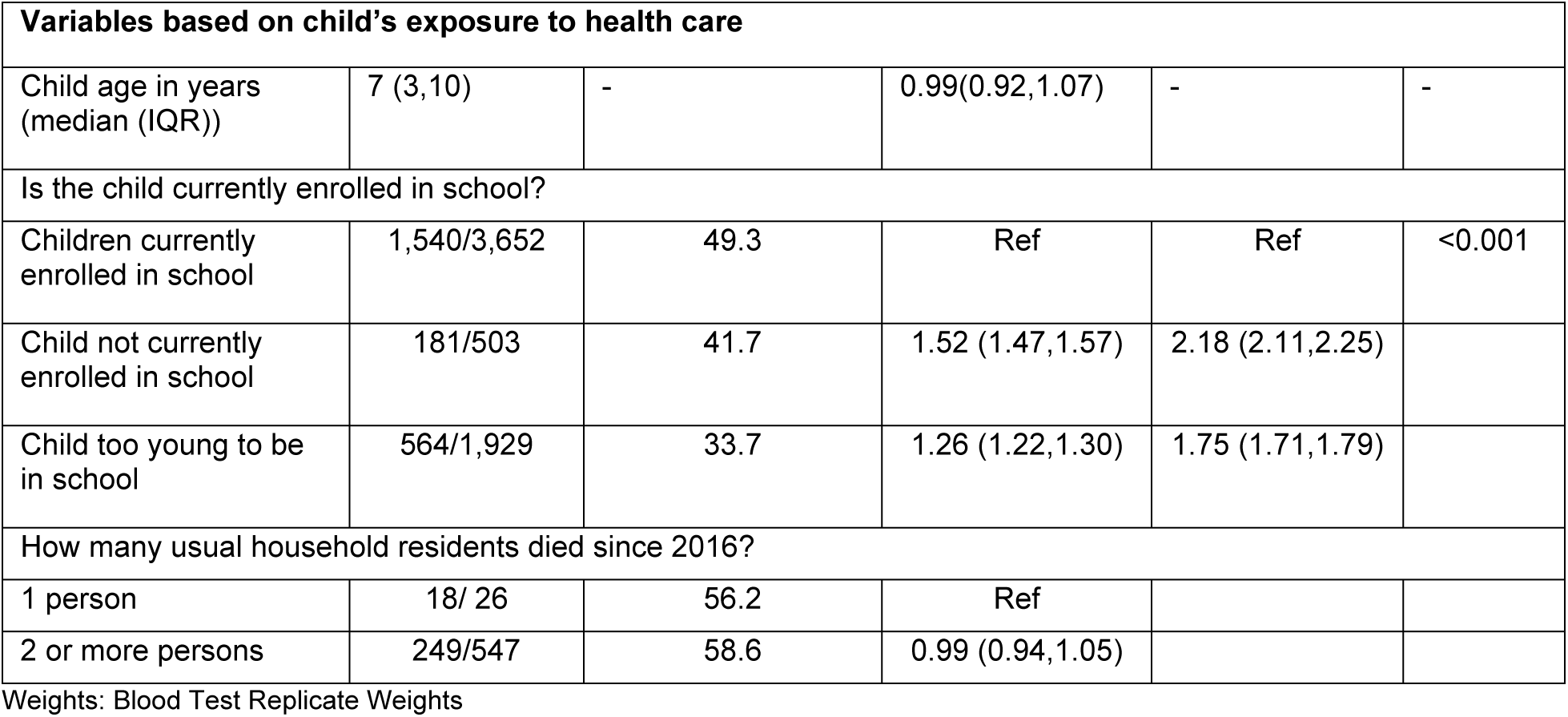
Factors associated with lack of HIV testing among children of women living with HIV and aware of their status, PHIA surveys from 13 countries, 2015 – 2019.

Overall, HIV treatment cascade was far below UNAIDS recommended targets of 95 – 95 – 95. Overall, of the 387 HIV-positive children in this survey, 74.6% (296) had a known HIV-positive status prior to the survey, 92.8% were on ART and 43.9% were virally suppressed (75-93-44). (Table 3).

**Table 3.** HIV treatment cascade of children of WLHIV and aware of their HIV positive status by age category, who tested HIV positive during the PHIA surveys from 13 countries, 2015 – 2019.

| Age category (years) | Children of WLHIV and aware of HIV status, with HIV positive test results during the survey, n (weighted col %) | Children known to be HIV-positive prior to the survey (weighted %) | Children who were on ART (weighted %) | Children who were suppressed (weighted %) |
| --- | --- | --- | --- | --- |
| <5 | 92 (31.9) | 56/92 (68.7) | 53/56(97.6) | 24/53 (27.7) |
| 5- 9 | 137 (40.0) | 109/137 (78.2) | 97/109(86.3) | 63/97 (49.0) |
| 10-14 | 158 (28.1) | 131/158 (76.1) | 129/131(97.2) | 90/129 (53.6) |
| Total | 387(100.0) | 296/387 (74.6) | 279/296(92.8) | 177/279 (43.9) |

\*Weights: Blood Test Replicate Weights

### Extrapolation

Overall, 2.1% (108/5,221) children living with HIV out of all children tested were not on ART during the time of the survey either due to previous unknown HIV-positive status or were aware of their HIV-positive status but not on ART. Overall the proportion of children newly diagnosed with HIV was 1.7%; the denominator excludes HIV positives with detectable ARVs. This extrapolates to 55,985 (95% CI: 55,972-55,998) children of HIV-positive women who were aware of their HIV status in the countries that participated in PHIA surveys, who had undiagnosed HIV or were HIV-positive but not on ART (Table 4).

**Table 4:** Estimated number of children living with HIV by diagnosis status among women with known HIV positive status, PHIA surveys from 13 countries 2015 – 2019.

| Age in years | Estimates of total children living with HIV<sup>1</sup> | Estimates of diagnosed children living with HIV<sup>2</sup> | Estimates of undiagnosed children living with HIV<sup>3</sup> |
| --- | --- | --- | --- |
| Overall | 218,613 (218,576 – 218,651) | 162,628 (162,603-162,653) | 55,985 (55,972-55,998) |
| 1–4 | 63,244 (63,225-63,261) | 41,942 (41,931-41,951) | 21,302 (21,294-21,310) |
| 5–9 | 88,557 (88,531-88,582) | 69,349(69,332-69,366) | 19,208 (19,198-19,217) |
| 10-14 | 66,812(66,796-66,827) | 51,337 (51,324-51,350) | 15,475 (15,472-15,477) |

<sup>1</sup> Tested HIV-positive in the PHIA
<sup>2</sup> Tested HIV-positive in the PHIA and were reported as positive by the parent/guardian or had detectable ARVs
<sup>3</sup> Tested HIV-positive in the PHIA but parent/guardian reported unknown HIV status or HIV-negative and child did not have detectable ARVs
Weights used: Blood Test Replicate Weights

## Discussion

We analyzed data from 13 nationally representative HIV-focused household surveys conducted in sub-Saharan Africa to determine the utility of family-based pediatric index testing. We found that slightly more than half of children were reported to have been tested for HIV prior to the survey. Our results were similar to previous reports of children of WLHIV who had been tested and ranged between 35% to 58% [8, 9, 15].

Among children with no detectable ARVs, we found that 2.9% of children that had never been tested and 0.9% reported to be HIV-negative were newly diagnosed. Previous studies have shown the test positivity among children of HIV-positive index clients varies by country and range from 1.9% to 23.0% [5, 16, 17]. The B-Gap study in Zimbabwe, focusing on HIV testing of children aged 2-18 years born to HIV-positive index clients who either had unknown HIV status or had tested HIV-negative more than 6 months ago found HIV prevalence to be 1.1% [16].

In our study, not visiting a health care worker in the last 12 months, absence of parental illness, non–disclosure of mother’s HIV status, rural residence, maternal high viral load and school attendance were significantly associated with the child never having been tested for HIV. Similar to our study, lack of information on the importance of testing, non-disclosure of maternal HIV status, having a deceased mother, urban residence, being asymptomatic and difficulties attending health facilities with children have been found to be associated with children of WLHIV not being tested [3, 5, 16, 18]. Other factors that have been reported to be associated with children of CLHIV not being tested include separation of children from parents, desire to involve mothers in the decision to test children, health care worker hesitancy to test and perceived unsuitability of accompanying parent/guardian to give consent for testing B-Gap study [17, 19]. Additional factors include attending clinic with a male parent/guardian, drug pickups for children in differentiated service delivery models and logistic challenges with contact tracing [5, 18, 19]. This analysis shows that on average, two children were linked and presumed elicited from every one woman living with HIV. This number may have been suboptimal as overall average births per woman of reproductive age as of 2020 in the focus countries ranged from 2.8 for Eswatini to 4.7 for Tanzania [20]. At the facility level, suboptimal elicitation of children of HIV-positive index clients is a major concern as this could result in missed opportunities to identify CLHIV. Routinely collected program data from adult female index clients aged 15 to 49 years, who accepted index testing in PEPFAR-supported programs in Malawi, Zimbabwe and Tanzania between April and June 2022, found the average number of children contacts identified per woman living with HIV to be 0.5, 0.7, and 1.2 respectively.

In this study, 9.0% of children known to be HIV-positive and tested HIV-positive, did not have detectable ARVs. These could be children who never initiated treatment or were off ART at the time of the survey. Distance from health facilities and poor relationship with health care workers may contribute to delayed ART initiation [21]. Documented factors affecting medication adherence in children include unavailability of caregivers, caregiver knowledge of ART, caregiver occupation and use of memory aids among others [22–24]. Up to 60% of children on ART may be reported as lost to follow up by 60 months after ART initiation [25, 26]. Some of these children may be documented as newly diagnosed if they present at different facilities.

We found that 82% of children who were not on ART and 62% of estimated number of CLHIV but undiagnosed were aged 5 years or older. These may be due to missed opportunities to prevent vertical HIV transmission or timely identification of CLHIV. Failure to repeat HIV testing of pregnant and breastfeeding women especially in countries with generalized HIV epidemics may contribute to missed opportunities to prevent vertical transmission and timely identification of CLHIV [4]. The risk of vertical transmission of HIV among women who seroconvert during pregnancy and breastfeeding period may be up to 19% [27]. Gaps in HIV case identification may be affected by lack of integration of HIV testing in maternal and child health clinics as well as community programs. In Kenya, HIV positivity among children of HIV-positive index clients tested in the community settings was 6.8% compared to 2.7% for those tested in health facilities [28].

We also found that 39.0% of those reported to have unknown HIV status, but tested HIV positive had detectable ARVs. Non-disclosure of HIV positive status may be due to stigma-related challenges around HIV positive diagnosis for the children and their mothers and possibly by extension their fathers. A systematic review that reported proportions of non-naïve adults initiating ART in sub-Saharan Africa found the proportions with evidence of prior ART use to be higher at 53% based on presence of ARV compared to self-report at 5% [29]. In Zambia the cumulative loss to follow up of children < 15 years of age initiated on ART was 16% and of the 90% traced, 13% were on ART at other health facilities while 14% continued ART with gaps. Some of these children may have presented as new clients without disclosing their HIV and ART status [30]. Gaps in data collection processes may also be another reason.

This study found 5.5% of children reported by their parent/guardian to be HIV positive tested HIV negative during the survey. This was unexpected given that WHO recommends repeat HIV testing to confirm status prior to release of HIV test results [4]. A few studies have reported the loss of detectability of HIV leading to ‘false-negative’ HIV-1 PCR results in a small proportion of children who start treatment in the first months of life and maintain good adherence [31, 32]. In a South African study, 2.2% of HIV-positive children initiated on ART at a median age of 6.3 months and had been taking ART for almost 10 years tested PCR negative [33]. PHIA surveys found that among adults aged 15 – 59 years who reported HIV-positive status during interview, 1.3%-1.4% tested HIV negative [34, 35]. Misunderstanding of the study questionnaire resulting in a wrong response may be another possibility. We also found that 0.9% of children who were reported as HIV negative by parent/guardians tested HIV-positive during the survey. This may have been a misreporting by the caregiver, who may not have been the biological parent. This may also have been the case in scenarios of non-disclosure and stigma-related consequences of an HIV positive diagnosis. It is also possible that some of these cases may result from vertically transmitted HIV from mothers who tested HIV negative during antenatal care but acquired HIV during breastfeeding period but did not receive interventions due to failure to retest. The proportion of mothers who test HIV positive following a previous HIV negative test result range between 0.3% to 1.2% [27, 36]. Although current HIV diagnostic tests have relatively high sensitivity and specificity, false positive and negative results may occur based on positive predictive value of the tests and HIV prevalence, and physicians have been advised to be aware of the specific causes of inaccurate results [37].

Our study had several limitations. The proportion of children with unknown HIV status may have been underestimated as only those whose mother were alive and aware of their HIV positive status during the survey period were included. Data on ARV detected was missing in 3.4% of children who tested HIV positive, and analysis assumed ARVs were not detected. This was also a quantitative study and therefore may not provide in-depth understanding of other contributing factors for missed opportunities for diagnosis of HIV-positive children. The PHIA surveys were conducted in different periods hence a possibility that participating countries implemented the survey under varied HIV policy guidelines and programmatic activities that may have affected access to HIV testing and ART for children. This analysis may have missed results from other PHIAs done after 2019 that may have included improved PVT interventions and pediatric HIV case identification.

The study also heavily depended on parent/guardian-reported unverified information on HIV status of the children before the survey and response rate was not 100%.

### Conclusion

Our findings show the importance of addressing gaps in family - based index testing and underscores the importance of ongoing efforts in pediatric HIV case identification and linkage to treatment for all CLHIV.

## Disclaimer

The findings and conclusions in this article are those of the authors and do not necessarily represent the official position of the funding agencies.

## PEPFAR attribution of support

This manuscript has been supported by the President’s Emergency Plan for AIDS Relief (PEPFAR) through the Centers for Disease Control and Prevention (CDC) under the terms of 13 cooperative agreements (Annex 1)

## Data Availability

PHIA data used for this analysis is publicly available

## Acknowledgements

We would like to acknowledge PEPFAR country teams, Ministries of Health of the participating countries and Centers for Disease Control and Prevention for technical support. We thank the data collection teams in the 13 counties as well as the participants who made this study possible

